# Aerobic Exercise Training Elevates Circulating sRAGE via Modulation of Sheddase Regulation in Adults with Type 2 Diabetes

**DOI:** 10.1101/2025.09.10.25335517

**Authors:** Ryan K. Perkins, Corey E Mazo, James Shadiow, Pallavi Varshney, Edwin R. Miranda, Victoria R. Miranda, Maggie Eisenberg, Elif A. Oral, Jacob M. Haus

## Abstract

**Objective:** Soluble receptors for advanced glycation end-products (sRAGE) and Toll-like receptor 4 (sTLR4) function as decoy receptors that blunt proinflammatory signaling in metabolic disease. This study examined the effects of 12 weeks of aerobic exercise (AE) training on circulating sRAGE, sTLR4, and associated skeletal muscle sheddase regulators in adults with type 2 diabetes mellitus (T2DM).

**Methods:** Thirty-three sedentary adults with T2DM (age 40–75 yrs) were randomized to AE (n=20) or control (CON; n=13). AE participants completed supervised aerobic training (60 min/day, 5 days/week) at 70% VO₂peak. Pre- and post-intervention assessments included oral glucose tolerance testing (OGTT), DXA, VO₂peak, and a standardized acute aerobic exercise trial with blood and skeletal muscle sampling. Circulating sRAGE, sTLR4, hsCRP, and skeletal muscle expression of TLR2/4, RAGE, MMP2/9, ADAM10, and TIMP1/3 were quantified.

**Results:** AE increased circulating sRAGE by +25.7% (P=0.005), with 83% of participants demonstrating a positive response. hsCRP decreased (−6.5%, P=0.04), while sTLR4 remained unchanged. AE also improved VO₂peak (+6.9%, P=0.001) and reduced body fat (−3.8%, P=0.00002). In skeletal muscle, TLR4 protein approached significance (−27.4%, P=0.10), and the TIMP3:ADAM10 ratio declined markedly (−87%, P<0.000001), suggesting reduced sheddase inhibition. No significant changes were observed in ADAM10 activity or MMP/TLR protein levels. sRAGE changes correlated with MMP9 expression (r=0.747, P=0.013).

**Conclusions:** AE training increases circulating sRAGE and improves systemic inflammatory profiles in adults with T2DM, potentially via modulation of sheddase regulatory balance. These findings identify AE training as a non-pharmacological strategy to enhance endogenous anti-inflammatory defenses in T2DM through sRAGE regulation.

## INTRODUCTION

Type 2 diabetes mellitus (T2DM) is a chronic metabolic disorder characterized by dysregulated glucose metabolism, leading to an increased risk of cardiovascular disease, kidney dysfunction, and other comorbidities (1). Lifestyle interventions, particularly aerobic exercise training, have demonstrated benefits for improving glycemic control and reducing the risk of complications associated with T2DM (2–5). Conserved inflammatory signaling pathways mediated by pattern recognition receptors (PRRs), including the receptor for advanced glycation end-products (RAGE) and Toll-like receptors (TLRs), have gained attention as key players in the pathogenesis of T2DM via chronic inflammation (6). RAGE, a multiligand PRR, is implicated in the cellular response to advanced glycation end-products (AGEs) and other pathogen- and damage-associated molecular patterns (PAMPs and DAMPs, respectively), perpetuating the inflammatory environment characteristic of T2DM. Similar to RAGE, TLR2 and TLR4 play fundamental roles in the innate immune response due to their overlap in activation by PAMPs and DAMPs, typically elevated in those with metabolic disease and convergence on downstream signaling mechanisms (7–10). Of the 10 TLRs in human skeletal muscle, TLR2 and TLR4 are among the most highly expressed (11, 12) and are also upregulated in those with metabolic dysfunction (13–15). It is through the inflammatory link, at least in part, that genetic deletion and blockade of RAGE (16–18) and TLR (19–21) signaling rescues metabolic function.

Extracellular matrix sheddases, including MMPs (matrix metalloproteinases) and ADAM10 (a disintegrin and metalloproteinase 10) and their negative regulators, such as tissue inhibitors of metalloproteinases (TIMPs) have well-established roles in the modulation of tissue remodeling (22). MMP2, MMP9, and ADAM10 are central to extracellular matrix (ECM) degradation and the shedding of PRRs, interrupting their downstream inflammatory effects. Proteolytic cleavage of PRR ectodomains via sheddase activity releases a solubilized receptor isoform, thereby appearing in cell-culture media or the circulation *in vivo* (*23–26*). Mechanistically, ADAM10 activity appears to be regulated by calcium (i.e., calcium calmodulin kinase, CaMK) and antidiuretic hormone (ADH)-induced G-protein coupled receptor signaling (25). Similarly, MMP2 and MMP9 are activated from their inactive pro forms under hypoxic conditions (27) and in the presence of growth factors (28). Exercise is known to greatly increase calcium availably for muscle contraction, reduce oxygen availability, and promote hormone-induced cell signaling events. Not surprisingly, several studies have illustrated that aerobic exercise (AE) and simulated exercise augments circulating sTLRs and sRAGE levels (7, 29–35) that have been shown to commonly be depressed in individuals with obesity and metabolic disorders (36–39). Animal studies have created a clear link between modifying circulating sRAGE concentration and prevention obesity-related inflammation and metabolic dysfunction (18, 40, 41). Acting as a decoy receptor for circulating PAMPs and DAMPs, sRAGE levels are typically regularly negatively correlated with obesity and glucose intolerance in humans (37, 42–47), providing insight into a potential therapeutic target.

Whether aerobic exercise regulates circulating sRAGE through changes in sheddase activity and their tissue inhibitors remains unknown.This study aimed to examine the impact of a supervised 12-week AE intervention on circulating sRAGE and sTLR4, as well as their regulatory components in individuals with T2DM. We also explored the effects of an acute AE trial on the circulating and skeletal muscle inflammatory environment involving RAGE, TLR2, and TLR4. We hypothesized that AE training would increase sRAGE levels and this observation would be related to changes in key MMPs and ADAM10, along with in their TIMP inhibitors.

## MATERIALS AND METHODS

### Participants

Female and male participants between the ages of 40-75 yrs were recruited from the greater Ann Arbor, MI area through multiple means, including physical flyers, social media, University of Michigan Medicine research databases, and word-of-mouth. Inclusion criteria consisted of current T2DM diagnosis, BMI of 26-44 kg/m^2^, and sedentary behavior (<30min of exercise, 3 times per week). Exclusion criteria included history of cardiovascular, cerebrovascular, renal or hematological disease, age <40 or >75 years, current tobacco use, and currently pregnant individuals. All participants provided informed, written consent before participation. This study was approved by the Institutional Review Board at the University of Michigan (#HUM00146001) and is registered at ClinicalTrials.gov (NCT03534687).

### Study Design

#### Randomization and Experimental Design

This study consisted of four study visits completed by all participants before and repeated after the intervention period. Briefly, visits included 1) consenting, screening, and oral glucose tolerance test (OGTT), 2) DXA body composition assessment, 3) VO2peak treadmill testing, and 4) acute AE treadmill trial. After completing physical activity and medical health history questionnaires during screening to determine eligibility, participants were randomized to a control (CON) or aerobic exercise training (AE) group (Figure 1) between February 2019 to May 2023. Randomization was performed using a 1:1 grouping allocation by a member of the research team with no clinical involvement in the trial. Participants randomized to CON were provided the opportunity to reenroll into the AE intervention upon completion of their original CON assignment. The intent of this reenrollment option was to provide equitable access to the intervention hypothesized to improve study outcomes and maximize adherence to preexisting lifestyle habits during the trial period, thereby reducing potential confounding factors.

**Figure 1.**
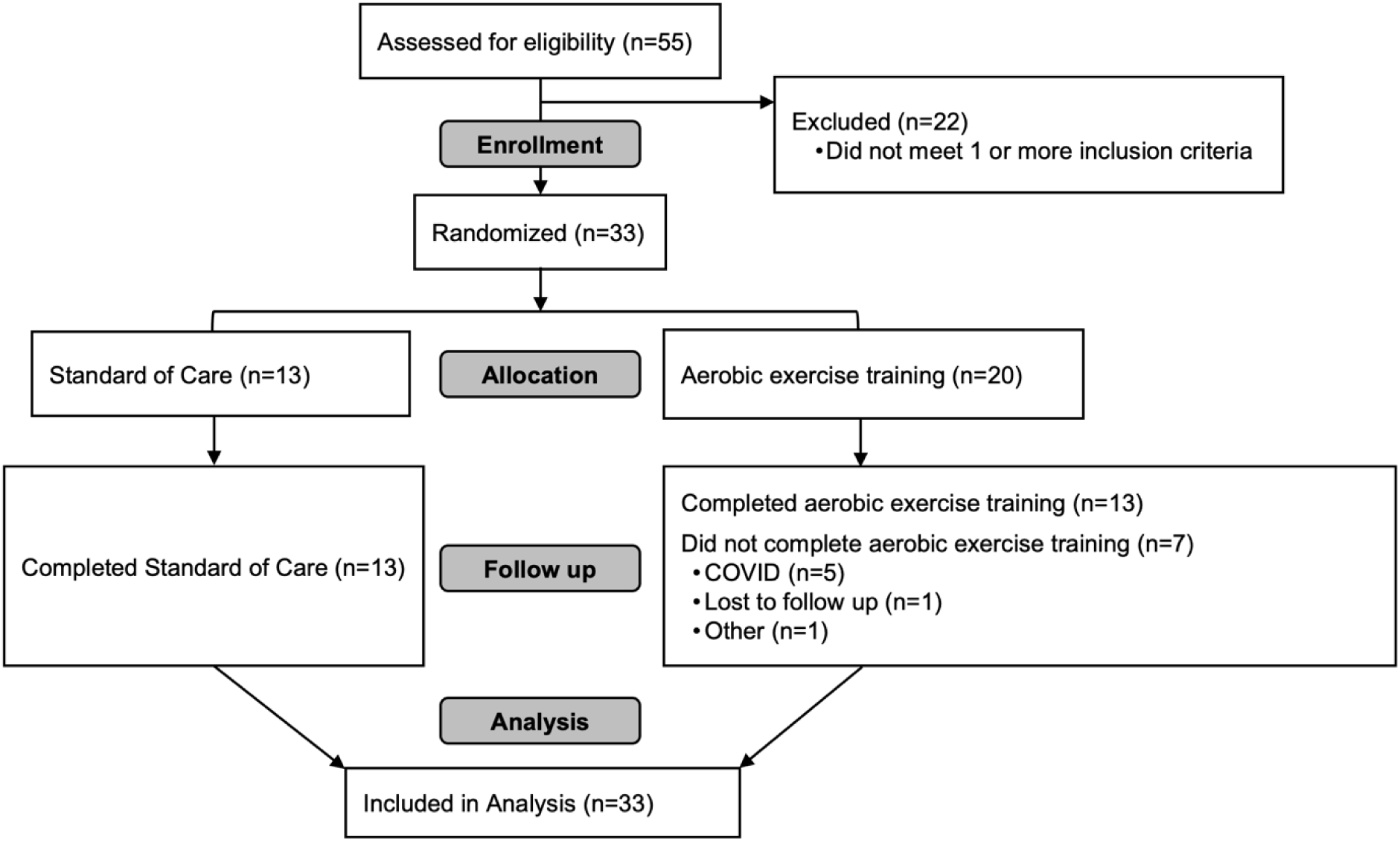
CONSORT Diagram. A total of 55 males and females underwent screening based on inclusion and exclusion criteria. Following screening, 33 participants were eligible to enroll in the 12-week intervention. Participants were randomized to an aerobic exercise training (n=20) or standard of care control group (n=13). 7 participants did not complete the AE training period. Final analysis was performed on 33 participants.

**Figure 2.**
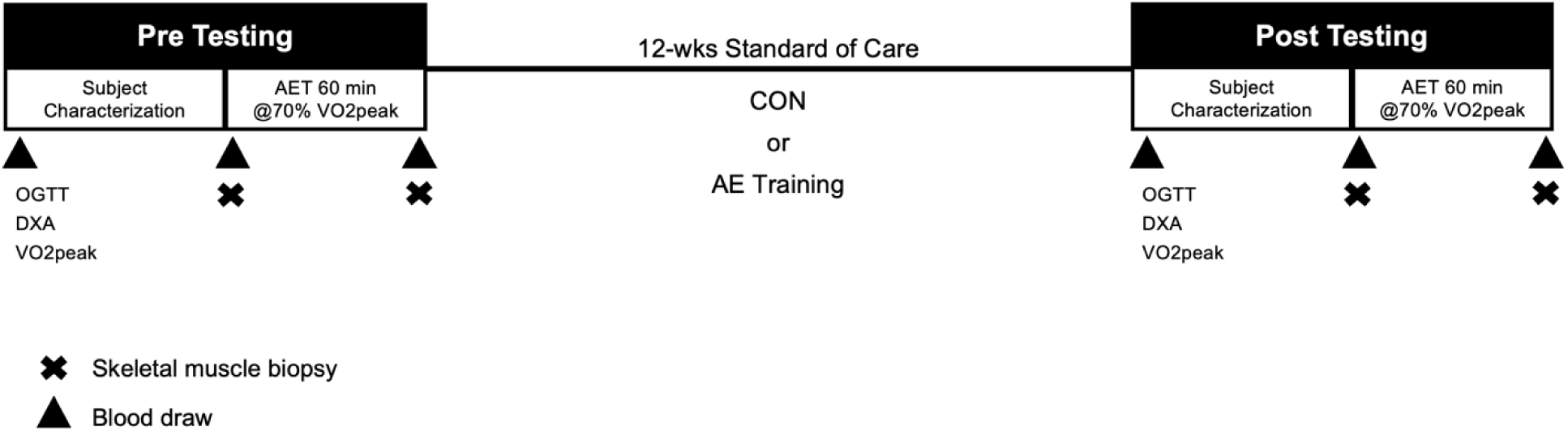
Prior to group randomization (CON, control; AE, aerobic exercise training), participants underwent body composition assessing (DXA), oral glucose tolerance testing (OGTT), and a VO2peak cardiovascular fitness test. Within 48 hr of randomization, participants underwent and a pre-intervention acute exercise trial (AET) for 60min at 70% VO2peak, with vastus lateralis skeletal muscle biopsies and blood draws taken before and 30-min post-AET. Following this pre-testing period, the CON group received 12-weeks of Standard of Care, while the AE group received 12-weeks Standard of Care and supervised aerobic exercise training. DXA, OGTT, VO2peak testing, and the AET was repeated following the 12-week intervention period.

A total of 55 participants expressed interest in the study, of which 22 were excluded due to inclusion and/or exclusion criteria (Figure 1). Therefore, 33 participants were randomized to either CON (n=13; 69% female) or AE (n=20; 70% female). Eight participants (75% female) originally randomized to CON elected to reenroll in AE after completing their original group allocation. In total, 7 participants did not complete the AE intervention period (i.e., lost-to-follow up, COVID logistical complications, or illness).

All participants were provided with American Diabetes Association Standard of Care for diabetes management (48) and were instructed to maintain current physical activity or diet habits outside of study-related activities. Individuals randomized to AE participated in 12-wks of supervised training sessions, consisting of treadmill, stationary cycle ergometer, elliptical, and/or upper-arm ergometry exercise. Initial VO2peak testing was conducted for all participants and training intensity for the AE group was based on heart rate (HR) responses. HR was monitored continuously during training sessions (Polar, Kempele, Finland) and intensity was adjusted to reach a HR that corresponded to the target % of VO2peak. Participants in the AE group exercised 5 days/week, with each session incorporating a short warmup and cooldown during training sessions to reach a total target duration of 60 min. The training program started with a 2-week ramp-up, familiarization period of progressing exercise intensity and duration. Exercise intensity began at a target intensity of 55% VO2peak for week 1 (40 min/session), progressing to 60-65% VO2peak for week 2 (50 min/session), and 70% VO2peak for all other weeks (60 min/session). Follow-up VO2peak tests were performed at weeks 4 and 8 to monitor progress and AE intensity was adjusted to match prescribed training HR (49). AE sessions were supervised by at least one member of the research team and all participants in AE completed >90% to exercise training sessions.

#### Metabolic control

For assessments sensitive to diet, drugs, and physical activity, standard metabolic controls were closely monitored. More specifically, before the OGTT and AET, participants were asked to refrain from eating for ∼10 hr (overnight fast), withhold oral antidiabetic medications the morning of testing, not consume alcohol for 48 hr, and abstain from physical activity and caffeine consumption for 24 hr prior to testing. To account for blood volume shifting that occurs with changes in body position or engagement in exercise (50), participants rested quietly (seated) for 30 minutes prior to all blood draws.

#### Visit 1 (OGTT)

Following screening, a 75-g OGTT was performed. Briefly, an antecubital or hand venous catheter was inserted, and baseline blood was collected. Participants consumed 75 g anhydrous glucose dissolved in 10 oz water within ∼3 min. Following consumption of the glucose drink, venous blood was sampled at regular intervals for 2 h (30-, 60-, and 120-min post).

#### Visit 2 (DXA body composition)

Height and weight were measured via stadiometer and digital scale, respectively. Body composition was assessed via dual X-ray absorptiometry (DXA; Lunar iDXA; GE Healthcare, Madison, WI).

#### Visit 3 (VO2peak testing)

Maximal oxygen consumption (VO2peak) was determined using a Cornell treadmill protocol until subjects reached volitional fatigue (51). Expired air was analyzed via indirect calorimetry (COSMED). HR was continuously monitored via 12-lead electrocardiogram, and subjects provided a rating of perceived exertion (RPE) at the end of each 2-minute stage. VO2peak was achieved in participants that met at least two of the five following criteria: volitional fatigue, a plateau in VO2 despite an increasing workload, RPE >17, RER > 1.1, and a heart rate >85% age-predicted maximal HR.

#### Visit 4 (Acute AE Trial)

All participants completed a supervised acute AE trial before and after the 12-week intervention. Upon arrival, participants rested quietly for 30 min (seated), after which a blood draw (antecubital vein) and *m. vastus lateralis* (VL) skeletal muscle biopsy was performed. The acute exercise session consisted of 60 min treadmill walking at a HR that corresponded with ∼70% of pre training VO2peak. VO2 was monitored for 5 min in 15 min intervals and speed and/or incline was adjusted to maintain target VO2 (Parvo Medics, Sandy, UT). Following the acute AE trial, participants rested quietly for 30 min, after which a final blood draw and VL biopsy was performed. This exercise bout was selected to provide a robust metabolic and cardiovascular challenge that aligns with American College of Sports Medicine and Physical Activity Guidelines for Americans (52, 53).

#### Blood Sample Collection

Blood was collected into EDTA and lithium-heparin coated vacutainers for the OGTT and the acute AE trail. For serum, whole blood was allowed to clot at room temperature for 30 min and was subsequently centrifuged at 3,500 RPM for 10 min at 20°C. For plasma, blood was centrifuged at 3,000 RPM for 10 min at 4°C. Following centrifugation, serum and plasma was stored at −80°C until analysis.

#### Skeletal muscle biopsy

Skeletal muscle biopsies were taken from the VL before and 30 min after the acute AE trial. Pre- and postexercise biopsies were alternated between left and right leg. Before the biopsy, local anesthetic (2% lidocaine HCL) was administered followed by a small incision (∼0.5cm) at the biopsy site (34, 54, 55). A Bergstrom biopsy needle was inserted with suction, extracting ∼200mg of muscle tissue. Muscle tissue was cleared of all visible connective tissue and fat, blotted with gauze to remove blood, immediately flash frozen in liquid nitrogen, and stored at −80°C until further analysis.

#### Circulating metabolites

Circulating glucose was determined using a point-of-care device (Contour Next One, Bayer Healthcare, Mishawaka, IN). Insulin was assessed via enzyme-linked immunosorbent assay (ELISA; 90095, Crystal Chem, Elk Grove Village, IL). To explore glucose homeostasis, Matsuda Index (56), homeostasis model of assessment-insulin resistance (HOMA-IR) (57), glucose AUC using the trapezoidal calculation (58, 59), disposition index (60), and insulinogenic index (IDI) was calculated (61).

Fructosamine quantification was determined via colorimetric assay (ab228558, Abcam, Cambridge, UK) following manufacturer’s instructions. Average inter-assay variability was 11.2% and intra-assay variability was 10-12.3%. Nonesterified fatty acids (NEFA) were measured via enzymatic assay (Wako HR series, Fujifilm, Lexington, MA). C-peptide was quantified via ELISA (DICP00, R&D Systems, Minneapolis, MN).

#### Plasma inflammatory factors

sRAGE was quantified via sRAGE ELISA (DRG000, R&D Systems, Minneapolis, MN) according to manufacturer’s instructions. The average inter-assay and intra-assay variability was 3.3% and 2.5-3.9%, respectively. sTLR4 (MBS167606, My Biosource, San Deigo, CA) was assessed via ELISA. hsCRP was determined via ELISA (#80955, Crystal Chem, Elk Grove Village, IL).

#### Skeletal muscle protein quantification and enzymatic activity assay

Protein expression in skeletal muscle was quantified via Western blot analysis. Approximately 20 mg (wet weight) of frozen muscle tissue was homogenized by ceramic beads (lysing matrix D, FastPrep-24 homogenizer, MP Biomedical, Santa Ana, CA) in 20 volumes of ice-cold 1X Cell Lysis buffer (#9803, Cell Signaling Technology, Danvers, MA), 1X Protease Phosphatase Inhibitor (#5872, Cell Signaling Technology, Danvers, MA) and 25µM-MG132 (F1100, UBPBio, Dallas, TX). Total protein concentration was determined via BCA assay (Pierce Biotechnology, Rockford, IL). Equal protein (25 μg) was loaded on a 10% pre-cast Criterion TGX (BioRad, Hercules, CA) and resolved using SDS-PAGE, transferred to a nitrocellulose membrane, and blocked with Odyssey Blocking Buffer (LI-COR Biosciences, Lincoln, NE) in TBS for 1.5 hr at room temperature. Primary antibody (Supplemental Table 1) diluted in blocking buffer containing 0.1% Tween 20 incubation took place over night overnight, at 4°C, with gentle rocking. After overnight incubation, membranes were washed 5 times with 1X TBST at room temperature for 5 min with vigorous shaking. Fluorophore-conjugated secondary antibodies were prepared at 1:10,000 dilution in blocking buffer containing 0.1% Tween 20 and membranes were incubated in the dark for 1 hr at room temperature. Secondary antibody incubations membranes were followed by 5 consecutive 1X TBST washes at room temperature for 5 min with vigorous shaking. Immunofluorescence was visualized with a NIR system (Odyssey, LICOR Biosciences, Lincoln, NE) and quantified using LI-COR Image studio software v 5.2 (LICOR Biosciences, Lincoln, NE). Following protein of interest quantification, membranes were stained with Revert total protein stain for normalization.

ADAM10 activity was determined using a fluorometric ADAM10 activity assay (AS-72226, Anaspec, Freemont, CA). Muscle samples were homogenized in ice cold assay buffer according to manufacturer’s instructions and 50 µg of protein was analyzed in duplicate (3915, Corning-Costar). Kinetic fluorescence was measured every 5 min for 60 min using SpectraMax ID5 Microplate Reader (Molecular Devices, Sunnyvale, CA).

#### Statistical Analysis

Statistical analyses were performed using PRISM (version 10.3.1, GraphPad Software, La Jolla, CA). Data normality was determined via Shapiro-Wilk test. Two-way repeated measures ANOVA or mixed effect model regression analyses (when time missing time points occurred) was used to explore differences between groups due to the intervention and responses to the AET. Bonferroni post hoc test was utilized to examine specific differences when appropriate. The Student’s two-tailed t-test was used to examine characteristic differences between groups at baseline and differences in responses between groups after the intervention period. Correlation analysis was assessed via Pearson correlation coefficient. Significance was accepted at P<0.05 and a trend was recognized when P>0.05 and P<0.10. Data are represented as mean ± SE with individual data points overlayed in figures.

## RESULTS

### Baseline subject characteristics

Baseline CON and AE group profiles are reported in Table 1. Groups were matched for age (P=0.13) and sex. Race was 88% white, 8% black and 4% Asian while ethnicity was 100% non-hispanic or latio. No differences were noted in HbA1c (P=0.13), circulating lipid profiles (total cholesterol: P=0.73, HDL: P=0.95, LDL: P=0.48, TG: P=0.68), TSH (P=0.91), Hb (P=0.50), and Hct (P=0.33) between groups at baseline.

**Table 1.** Baseline subject characteristics.

| Variable, units | CON | AE |
| --- | --- | --- |
| *n*, % female | 13, 69% | 20, 70% |
| Age, yr | 59.6 ± 2.6 | 54.5 ± 2.1 |
| Height, m | 1.68 ± .03 | 1.67 ± .02 |
| HbA1c, % | 6.9 ± 0.2 | 7.5 ± 0.3 |
| TSH ,mIU/l | 1.9 ± 0.3 | 2.0 ± 0.4 |
| Cholesterol, mg/dl | 166 ± 11 | 172 ± 11 |
| HDL, mg/dl | 45 ± 3 | 45 ± 3 |
| LDL, mg/dl | 88 ± 8 | 96 ± 9 |
| Triglycerides, mg/dl | 168 ± 28 | 153 ± 19 |
| Hb, g/dl | 14.1 ± 0.4 | 13.7 ± 0.4 |
| Hct, % | 43 ± 1 | 42 ± 1 |

TSH, thyroid stimulating hormone; HDL, high-density lipoprotein; LDL, low-density lipoprotein; Hb, hemoglobin; Hct, hematocrit. Group differences assessed via two-tailed t-test.

### Anthropometrics

Anthropometric subject characteristics are reported in Table 2. CON and AE did not differ in weight, BMI, lean mass, fat mass, or body fat % before the intervention. AE decreased fat mass (−4.4±1.3 %, P=0.01) and body fat % (−3.8±1.0 %, P=0.00002) following the 12-wk training program. Further, there was a differential change in lean mass (CON: −0.7±0.3, AE: +0.9±0.6 kg, P=0.02) between groups following the intervention while weight (group effect P=0.58, time effect P=0.67, group x time P=0.24). and BMI (group effect P=0.45, time effect P=0.96, group x time P=0.14) remained unchanged.

**Table 2.** Anthropometric, cardiovascular fitness, and glucose metabolism changes after the 12-week intervention.

| Variable, Units | <u>CON</u> |  | <u>AE</u> |  |
| --- | --- | --- | --- | --- |
|  | Pre | Post | Pre | Post |
| *Anthropometrics* |  |  |  |  |
| Weight, kg | 91.8 ± 5.2 | 92.4 ± 4.9 | 96.4 ± 4.2 | 91.3 ± 5.2 |
| BMI, kg/m<sup>2</sup> | 32.6 ± 1.6 | 33.3 ± 1.6 | 34.6 ± 1.3 | 32.9 ± 1.9 |
| Lean Mass, kg | 53.4 ± 3.2 | 55.6 ± 3.1 | 54.1 ± 2.3 | 55.1 ± 2.7 |
| Fat Mass, kg | 36.6 ± 3.3 | 39.3 ± 3.4 | 41.9 ± 2.6 | 36.1 ± 4.3* |
| Body Fat, % | 40.7 ± 2.1 | 41.4 ± 2.4 | 43.6 ± 1.5 | 38.7 ± 2.7** |
| *Cardiovascular Fitness* |  |  |  |  |
| VO<sub>2</sub>peak, ml/kg/min | 21.1 ± 1.0 | 21.5 ± 1.4 | 22.9 ± 1.0 | 26.3 ± 1.3** |
| VO<sub>2</sub>peak, L/min | 1.9 ± 0.1 | 2.0 ± 0.1 | 2.2 ± 0.1 | 2.4 ± 0.1* |
| *Glucose Metabolism* |  |  |  |  |
| FPG, mg/dL | 142.5 ± 8.6 | 145.8 ± 10.2 | 158.8 ± 9.8 | 142.5 ± 10.5 |
| OGTT Glucose 2h, mg/dL | 274.8 ± 21.2 | 260.0 ± 17.6 | 278.2 ± 14.8 | 255.4 ± 23.0 |
| OGTT AUC, mg/dL/hr | 13798 ± 1036 | 11698 ± 850 | 12946 ± 671 | 12020 ± 1111 |
| HOMA-IR | 4.2 ± 0.6 | 4.1 ± 0.7 | 5.3 ± 1.2 | 3.1 ± 0.6 |
| Matsuda Index | 3.3 ± 0.4 | 3.3 ± 0.4 | 3.5 ± 0.5 | 6.3 ± 1.6* |
| Disposition Index | 0.47 ± 0.08 | 1.32 ± 0.46 | 0.82 ± 0.29 | 1.21 ± 0.38 |
| Insulinogenic Index | 0.17 ± 0.04 | 0.45 ± 0.19 | 0.31 ± 0.12 | 0.37 ± 0.16 |
| Fructosamine (μM) | 430.6 ± 50.6 | 452.5 ± 38.1 | 488.9 ± 31.2 | 374.8 ± 32.9* |

BMI, body mass index; OGTT, oral glucose tolerance test; HOMA-IR, homeostatic model of insulin resistance; \*P<0.05 vs Pre; \*\*P≤0.001 vs Pre. Pre- and post-intervention variables were analyzed using mixed model regression analyses with Bonferroni correction for multiple comparisons.

**Table 3.**
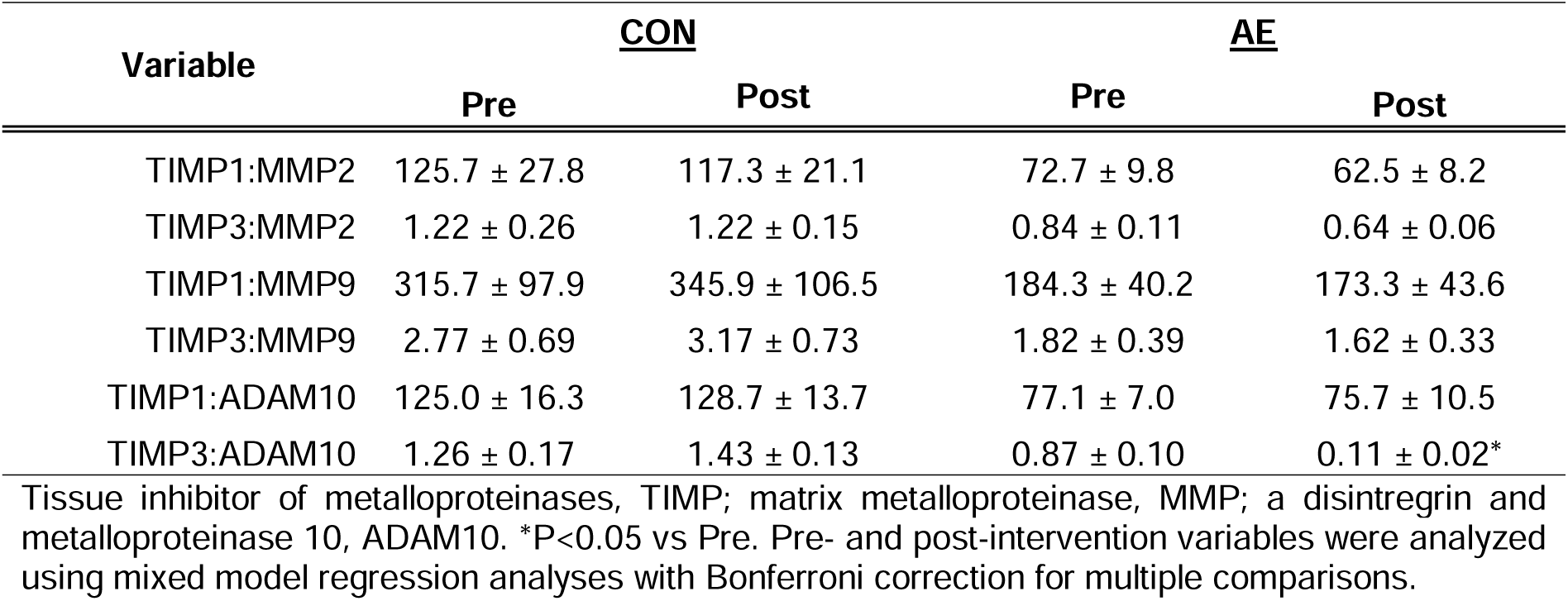
Relationships between TIMPs, MMPs, and ADAM10 before and after the 12-week intervention.

### Cardiovascular fitness

Relative and absolute VO2peak was similar (P>0.05) between CON and AE at baseline (Table 2). Relative VO2peak (+6.9±1.8 %, P=0.001) and absolute VO2peak (+5.0±1.9 %, P=0.03) increased in the AE group following the intervention period. O2pulse was similar between groups before the intervention (Pre: CON 12.1±0.9, AE 13.5±0.8 mL O2/bt; P=0.50) and increased in AE following the training period (Post: CON 12.3±1.0, AE 14.9±1.0 mL O2/bt; AE +6.1±2.5 %, P=0.03). HRpeak was not different between groups at baseline (P=0.99) and did not change following the intervention (data not shown).

### Glucose metabolism

All indices of glucose metabolism (Table 2) were similar between CON and AE at baseline (P>0.05). Following the 12-wk intervention, Matsuda Index increased (+78.7±53.7 %, P=0.02) and circulating fructosamine concentration decreased (−16.2±6.7 %, P=0.02) in AE. Circulating NEFA (μM) was similar between groups before the intervention (CON:415.4±33.0, AE: 537.6.7±62.0, P=0.15). Though not statistically significant, baseline NEFA decreased in AE following the intervention (−114.7±57.3 μM, −12.9±16.8%, P=0.13). FPG (group effect P=0.65, time effect P=0.30, group x time P=0.16), OGTT 2hr glucose (group effect P=0.86, time effect P=0.13, group x time P=0.45), OGTT glucose AUC (group effect P=0.59, time effect P=0.03 group x time P=0.71), HOMA-IR (group effect P=0.73, time effect P=0.29, group x time P=0.27), disposition index (group effect P=0.81, time effect P=0.04, group x time P=0.36), and insulinogenic index (group effect P=0.92, time effect P=0.13, group x time P=0.20) remained unchanged in both groups.

### Circulating inflammatory profile

Basal hsCRP was similar between groups at baseline and decreased in AE following the intervention (−6.5±21.2 %, P=0.04). CON and AE exhibited similar basal circulating sRAGE before the intervention (Figure 3A) and sRAGE increased in AE following the training period (+178±54 pg/mL, +25.7±9.3 %, P=0.005; Figure 3B, 3E). Of note, nearly all participants in AE increased basal sRAGE following exercise training (83 %). Following the acute aerobic exercise challenge, sRAGE did not change in CON or AE, before or after the intervention (Pre: CON −12.1±1.7, AE −11.9±2.6 %; Post: CON −13.6±2.8, AE −7.0±3.0 %; group effect P=0.21, time effect P=0.53, group x time P=0.23). Basal sTLR4 was similar between CON and AE before and after the intervention (Figure 3C) and did not change in response to the acute aerobic exercise challenge in either group (Pre intervention: CON +0.5±2.5, AE 4.1±4.3 %; Post intervention: CON +0.6±2.9, AE +9.9±7.2 %; group effect P=0.17, time effect P=0.53, group x time P=0.54; Figure 3D-E).

**Figure 3.**
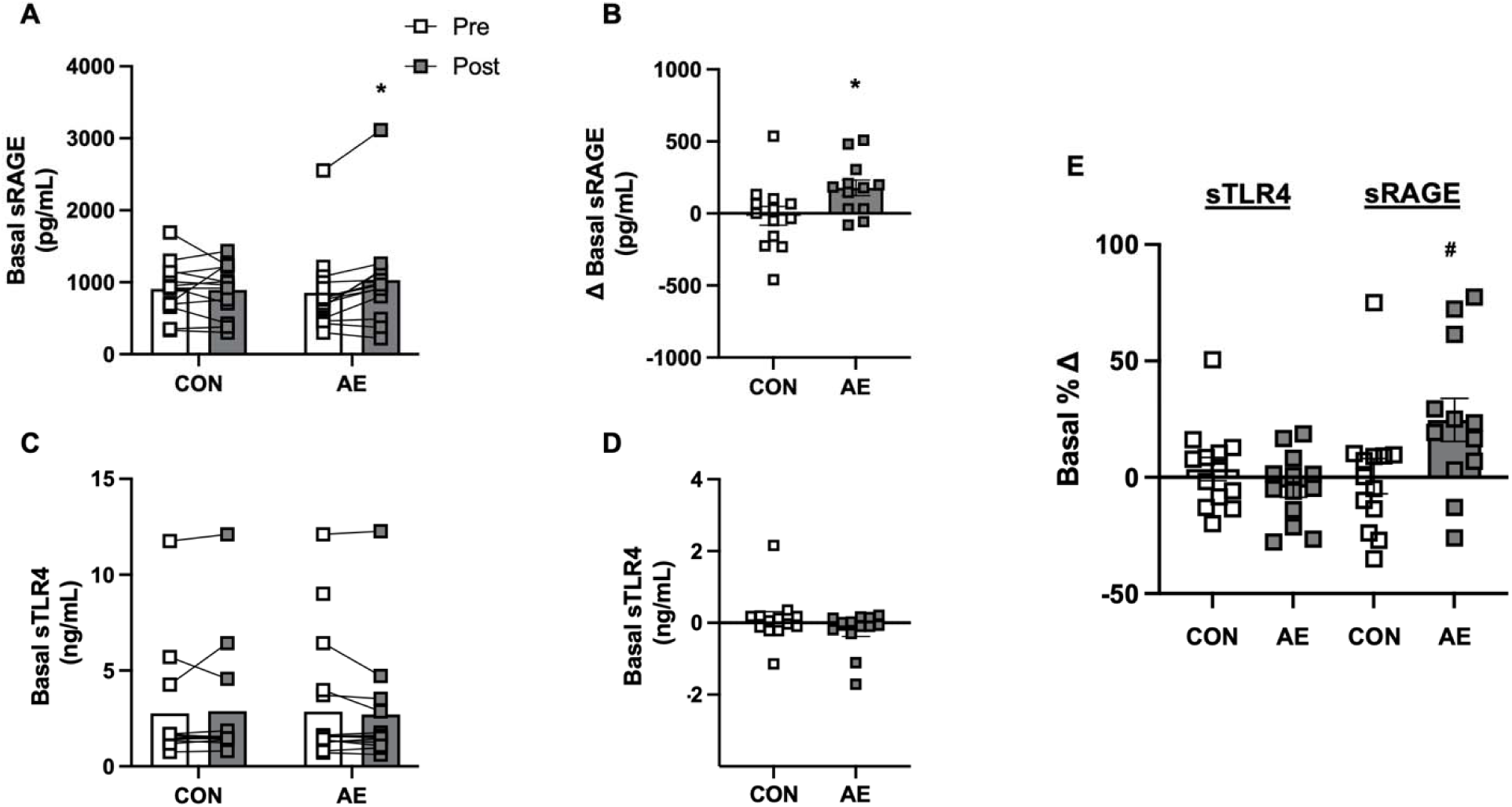
Basal solubilized receptor for advanced glycation end products (sRAGE; A), solubilized Toll-like receptor 4 (sTLR4, C) pre and post 12-week intervention in the control (CON) and aerobic exercise training (AE) groups. Respective absolute changes in basal sRAGE (B) and sTLR4 (C) and percentage changes in sRAGE (E) and sTLR4 (E) were calculated. *P≤0.05 vs Pre, ^#^P≤0.05 vs CON. Statistical analyses performed include mixed model regression analyses with Bonferroni post hoc (panels A & C) and two-tailed t-test (panels B, D, & E).

### Skeletal muscle inflammatory profile

TLR4 protein expression was similar between groups at baseline and approached significance in the AE group following training (−27.4±10.0 %, P=0.10; Figure 4B). TLR2 (group effect P=0.18, time effect P=0.70, group x time P=0.18) and RAGE (group effect P=0.84, time effect P=0.52, group x time P=0.10) protein expression did not differ between groups or change following the intervention (Figure 4A, 4C). To characterize alterations in MMP composition, we evaluated protein expression of the pro and active forms of MMP-2 (pro: 72 kDa, active: 64 kDa), MMP-9 (pro: 92 kDa, active: 84 kDa), and ADAM10 (pro: 100 kDa, active: 60 kDa) (Figure 5A-F). Expression of MMP2’s active form showed a trend to increase after training in the AE group (+12.4±4.7 %, P=0.07). Expression of MMP2 (pro form only) MMP9 (pro and active form) and ADAM10 (pro and active form), and the ratios between their pro and active form (data not shown), was similar between groups and tended to change in CON (P=0.08, but not AE (P=0.56). Basal ADAM10 activity was similar between groups at baseline (P=0.99) and tended to increase in CON following the intervention (−12.5±3.5 %, P=0.08), but not AE (−1.3±5.2%,). Following the AET, ADAM10 activity did not change in either group before the intervention (CON: −2.1±6.3%, AE: −0.2±6.8%, P=0.56). Known MMP regulators, TIMP1 (group effect P=0.23, time effect P=0.19, group x time P=0.83) and TIMP3 (group effect P=0.42, time effect P=0.24, group x time P=0.07), were also similar between groups at baseline and remained unchanged following the intervention period (Figure 5H-I). Because MMP expression and function are sensitive to TIMP1 and TIMP3, we also sought to quantify potential changes in their ratios (TIMP:MMP active form) before and after the intervention (Figure 6). Following exercise training, the AE group greatly reduced basal expression of TIMP3:ADAM10 (−86.9±2.0 %, P<0.000001). In addition, there was a marked difference in the change in TIMP3:MMP9 between groups (26% difference, P=0.04). No other distinctions in TIMP:MMP were found (all P>0.05).

**Figure 4.**
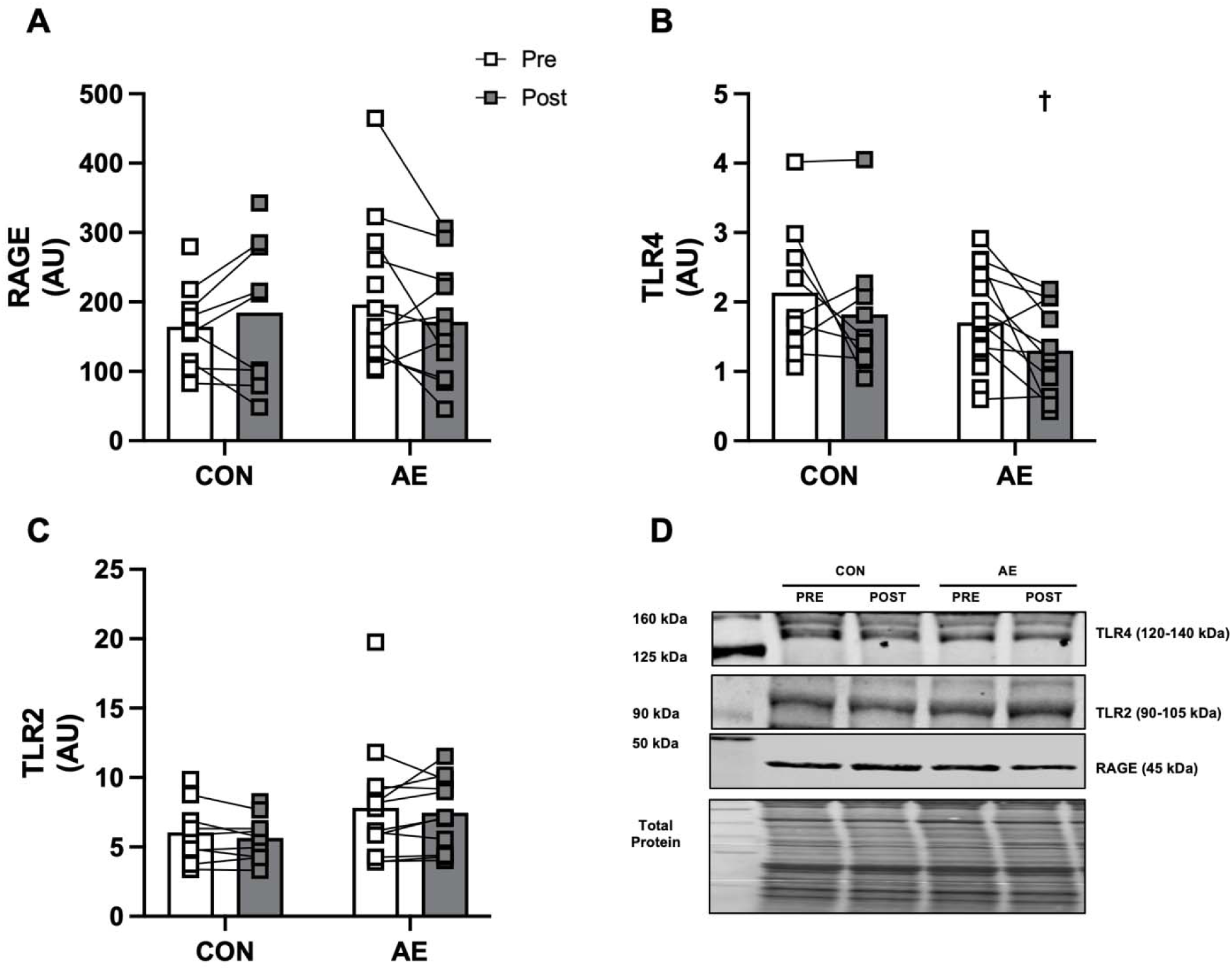
Basal vastus lateralis skeletal muscle receptor for advanced glycation end products (RAGE, A), Toll-like receptor 4 (TLR4, B), and Toll-like receptor 2 (TLR2, C) protein expression pre and post 12-week intervention in the control (CON) and aerobic exercise training (AE) groups. Western-blot quantification was normalized to total protein (D). ^†^P≤0.10 vs Pre. Statistical analyses performed include mixed model regression analyses with Bonferroni post hoc (panels A-C).

**Figure 5.**
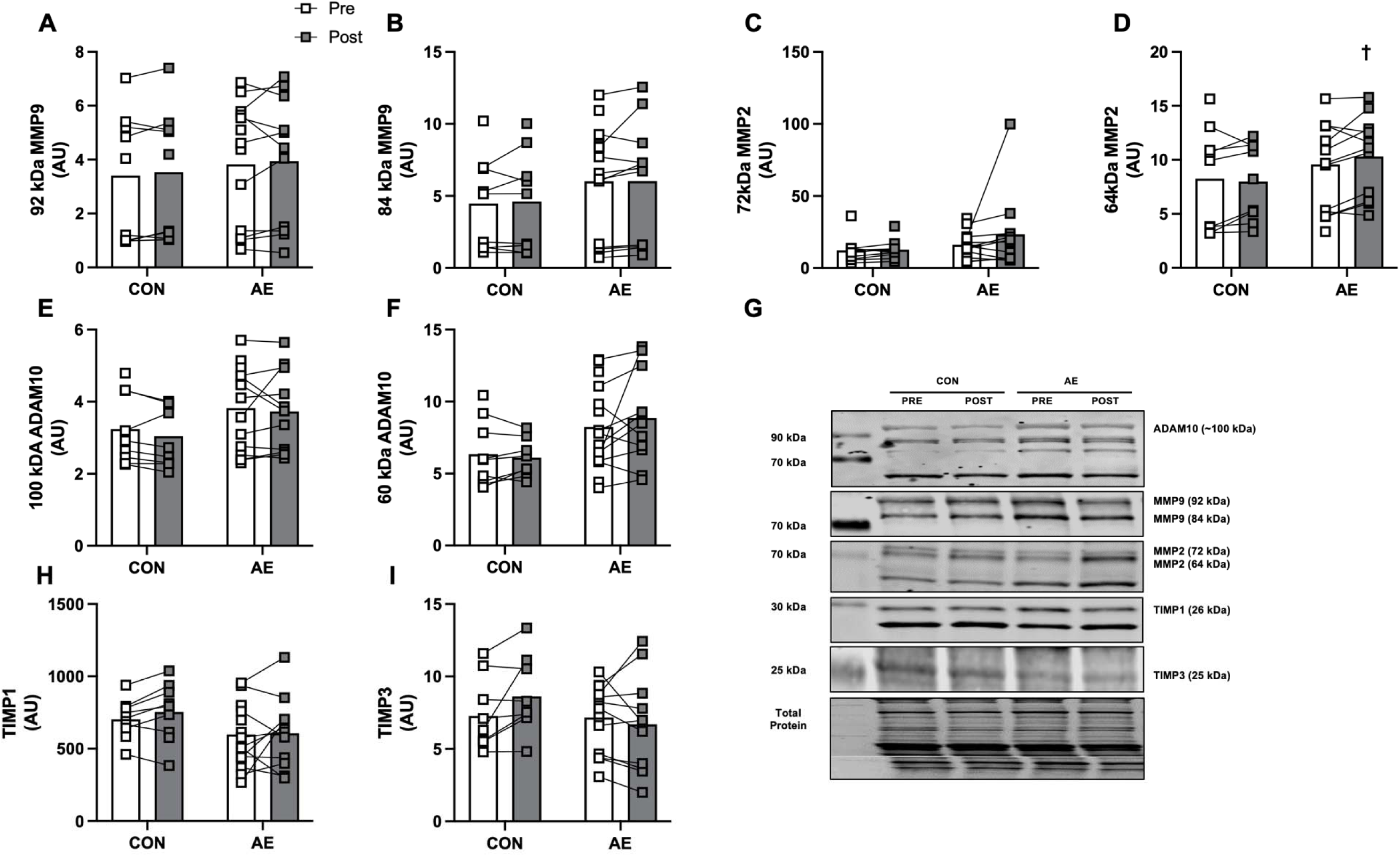
Basal vastus lateralis skeletal muscle pro (A) and active (B) matrix metalloproteinase (MMP) 9, pro (C) and active (D) MMP2, pro (E) and active (F) a disintegrin and metalloproteinase 10 (ADAM10), tissue inhibitors of metalloproteinases (TIMP) 1 (H) and TIMP3 (I) protein expression pre and post 12-week intervention in the control (CON) and aerobic exercise training (AE) groups. Western-blot quantification was normalized to total protein (G). ^†^P≤0.10 vs Pre. Statistical analyses performed include mixed model regression analyses with Bonferroni post hoc (panels A-I).

**Figure 6.**
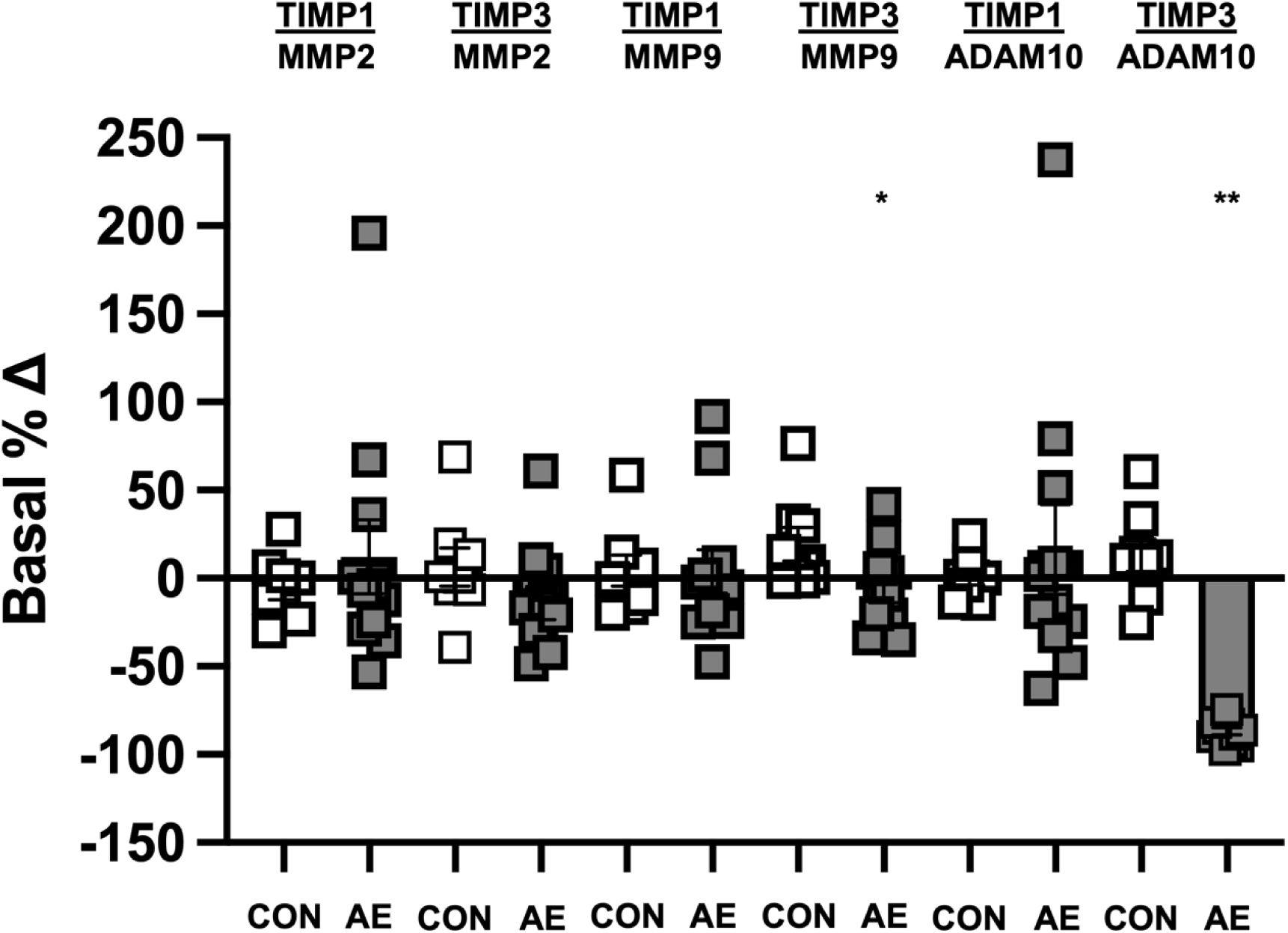
Percentage changes in basal expression ratios of matrix metalloproteinases (MMP2 and MMP9), a disintegrin and metalloproteinase 10 (ADAM10), and their known tissue inhibitors of metalloproteinases (TIMP1 and TIMP3). *P<0.05 vs CON. **P<0.001 vs CON). Statistical analyses performed include two-tailed t-test (panels B, D, & E).

### Notable relationships in AE

Several noteworthy relationships between changes in cardiovascular fitness and indices of glucose homeostasis and inflammatory factors were found (Table 4). For inflammatory factors and their known regulators following the training program, there was a relationship between the % changes in baseline TLR2 and MMP2 64 kDa (r=0.646, P=0.03), sRAGE and hsCRP (r=0.509, P=0.09), sRAGE and MMP9 92 kDa (r=0.747, P=0.01), sRAGE and MMP9 84/92 kDa (r=0.626, P=0.05), sTLR4 and ADAM10 100 kDa (r=0.649, P<0.05), sTLR4 and TIMP1 (r=-0.629, P<0.05), sTLR4 and MMP2 64 kDa (r=0.642, P=0.03).

**Table 4.** Select correlations between changes in cardiovascular fitness, indices of glucose metabolism, solubilized receptors and their known regulators after the 12-week intervention.

| Variable | Absolute<br>VO<sub>2</sub>peak<br>(%Δ) | Insulinogenic<br>Index<br>(%Δ) | Matsuda Index<br>(%Δ) | Disposition Index<br>(%Δ) | FPG<br>(%Δ) |
| --- | --- | --- | --- | --- | --- |
| sTLR4 | 0.001, P=0.99 | -0.295, P=0.35 | 0.543, P=0.07 | 0.409, P=0.19 | -0.570, P=0.04 |
| sRAGE | 0.231, P=0.47 | 0.675, P=0.02 | -0.047, P=0.89 | 0.167, P=0.60 | 0.375, P=0.23 |
| ADAM10 100 kDa | 0.425, P=0.19 | 0.400, P=0.25 | 0.771, P=0.01 | 0.849, P=0.002 | 0.306, P=0.35 |
| ADAM10 60 kDa | 0.361, P=0.27 | 0.592, P=0.07 | 0.095, P=0.79 | 0.271, P=0.45 | -0.159, P=0.64 |
| MMP2 72 kDa | -0.331, P=0.32 | -0.058, P=0.87 | -0.091, P=0.80 | -0.046, P=0.90 | 0.101, P=0.77 |
| MMP2 64 kDa | -0.053, P=0.88 | -0.018, P=0.96 | 0.150, P=0.68 | 0.101, P=0.78 | -0.290, P=0.39 |
| MMP9 92 kDa | 0.635, P=0.04 | 0.653, P=0.04 | 0.051, P=0.89 | 0.227, P=0.53 | -0.306, P=0.36 |
| MMP9 84 kDa | -0.152, P=0.65 | 0.096, P=0.79 | -0.224, P=0.53 | -0.135, P=0.71 | 0.385, P=0.24 |

Soluble Toll-like receptor 4, sTLR4; soluble receptor for advanced glycation end products, sRAGE; matrix metalloproteinase, MMP; a disintegrin and metalloproteinase 10, ADAM10. Relationships assessed via Pearson correlation coefficient.

## DISCUSSION

This study investigated the impact of a 12-week AE intervention on circulating sRAGE, sTLR4, and their associated regulators in individuals with T2DM. Additionally, we explored the effects of an acute AE trial on skeletal muscle inflammatory signaling, specifically targeting RAGE, TLR2, and TLR4. Our findings demonstrate that AE training significantly increased circulating sRAGE levels and decreased hsCRP levels, with accompanying changes in body composition, cardiovascular fitness, and some aspects of glucose metabolism. In general, these effects were not accompanied by substantial alterations in inflammatory markers related to extra-cellular matrix remodeling within skeletal muscle. Overall, these findings suggest a potential role for AE training in modulating systemic inflammation rather than directly influencing local mediators of muscle inflammation.

sRAGE is considered a decoy receptor that binds AGEs and other DAMPs and PAMPs, thereby mitigating their inflammatory effects (7, 31–33). In our study, AE training led to a 27% increase in circulating sRAGE as most of the participants responded to the training program (>80%). This finding is consistent with other reports indicating that AE can enhance sRAGE levels, potentially reducing chronic inflammation and metabolic dysfunction (29, 31–33). Notably, the ratio of pro- to active-form MMP9 showed a marked correlation with sRAGE levels following exercise training (r=0.626, P=0.05), providing valuable insight into the observed increase in sRAGE. While the acute AE trial did not result in a change in sRAGE levels, long-term AE led to a significant increase. This suggests that exercise-induced upregulation of sRAGE may be a sustained response that occurs outside of the 30-minute post-exercise period.

In contrast to sRAGE, sTLR4 levels were not significantly altered by either the 12-week AE intervention or the acute AE bout. This finding suggests that, while tissue-bound TLR4 plays a crucial role in innate immune responses and inflammation related to metabolism (7–9, 30), its regulation by exercise may differ from that of sRAGE. Though sTLR4 levels have been shown to be 54% lower in individuals with metabolic dysfunction (36) and exercise influences TLR4 expression in skeletal muscle and other tissues (7, 11, 30, 62), our results suggest that individuals with T2DM may be less sensitive to modification with AE. This discrepancy may be due to differences in the stimuli needed to, or the mechanisms that lead to the production of sRAGE versus sTLR4 with exercise.

A key aspect of the study was to examine whether exercise modulates the activity of MMPs and their negative regulators, TIMPs, particularly in relation to RAGE and TLR signaling. Previous exercise training studies show an increase in skeletal muscle MMP gene expression and activity with exercise in healthy individuals (63), but likely to a lesser degree than those with T2DM (64, 65). Although here we report no significant changes in the expression of MMPs (MMP2, MMP9) or ADAM10 due to AE training, we did observe a differential change in the ratio of TIMP3:MMP9 between groups. In addition, we found an overt decrease in the ratio of TIMP3:ADAM10 in the AE group, suggesting that AE may reduce the inhibitory regulation of ADAM10, potentially leading to more effective receptor shedding. A notable finding was the 87% reduction in TIMP3:ADAMP10 ratio, indicating reduced sheddase inhibition and aligning with previous studies showing that exercise can influence the balance of TIMP:MMP ratios (64). Moreover, our study found a pronounced correlation between changes in sRAGE and MMP9, indicating that exercise-induced increases in sRAGE might be linked to the activity of MMPs in the systemic circulation, possibly contributing to a reduction in chronic inflammation.

Interestingly, while protein expression of TLR2 and RAGE in skeletal muscle did not change substantially with AE, TLR4 expression tended to decrease in the AE group. This finding, although trending toward statistical significance, is noteworthy as TLR4 expression has been demonstrated to be elevated in individuals with metabolic disease, (13–15) and interrupting TLR4 signaling improves metabolic function (19–21). Because changes in TLR4 was not associated with other variables of interest (e.g., sTLR4, regulatory shedding factors), this suggests that exercise may have a localized impact on reduced TLR4 signaling in skeletal muscle, possibly via adaptations that diminish inflammatory signaling pathway activity that lead to its production (9). These results also suggest that while AE is known to have systemic anti-inflammatory effects, it may not necessarily influence local muscle inflammation to the same extent, possibly due to the nature of T2DM or the specific exercise protocol used in this study.

The relationship between changes in body composition, cardiovascular fitness, and inflammatory markers provides further insight into the mechanisms underlying AE’s effects on T2DM. Our finding that AE training reduced fat mass and body fat percentage is consistent with previous research indicating that exercise can markedly improve body composition in individuals with T2DM (29, 32). The positive changes in cardiovascular fitness reported here, indicated by increased VO2peak, aligns with well-established observations that exercise improves cardiovascular function and glucose metabolism in individuals with T2DM (66). Moreover, the observed correlations between changes in cardiovascular fitness and inflammatory factors, such as hsCRP and sRAGE, suggest that improved fitness levels may mediate some of the observed reductions in inflammation, independent of weight loss.

Despite improvements in the Matsuda Index and circulating fructosamine, the lack of substantial changes in other glucose metabolism indices (such as HOMA-IR, FPG, and OGTT AUC) after AE training, is an important consideration. These results suggest that while AE training can improve insulin sensitivity and glycemic control in T2DM, the effects may be more evident in some indices (e.g., Matsuda Index) compared to others (e.g., HOMA-IR). Moreover, while NEFA levels did not notably decrease in the AE group, there was a trend towards a reduction, suggesting that exercise may indirectly impact lipid metabolism and its association with inflammation, particularly as a known PAMP for TLR4-induced signaling (9).

While this study provides valuable insights into the effects of AE on inflammation and receptor shedding in T2DM, several limitations must be considered. The relatively short duration of the intervention (12 weeks) may have limited the ability to detect long-term changes in key inflammatory pathways and ECM remodeling processes. Additionally, the focus on AE may not capture the full spectrum of exercise-induced changes in inflammation and ECM remodeling, as resistance training or high-intensity interval training may have unique effects. Future studies should explore the effects of longer-duration exercise interventions and investigate whether different exercise modalities can further regulate inflammation and receptor shedding in T2DM. Furthermore, the underlying mechanisms linking changes in inflammatory markers and improvements in glucose metabolism remain to be fully elucidated, and additional studies are needed to explore the role of these markers in the development and progression of T2DM. Lastly, the source of circulating sRAGE and sTLRs in our study is unknown. Though skeletal muscle is a predominant tissue by size and metabolic activity in humans and these solubilized receptors have been reported to be muscle-derived, other cell types (i.e., adipose, vascular, and immune) may also contribute to the circulating sRAGE and sTLR pool and require further investigation.

Collectively, these findings underscore aerobic exercise as a promising adjunctive strategy to augment endogenous anti-inflammatory signaling in T2DM, with sRAGE serving as a potential biomarker of therapeutic response. The increase in circulating sRAGE, alongside reductions in hsCRP and changes in MMP:TIMP ratios, suggests that AE may modulate immune responses and tissue remodeling mechanisms, potentially contributing to improved metabolic health. Further studies are needed to explore the long-term effects of exercise on inflammatory pathways, the impact of different exercise modalities on muscle inflammation, and the potential for combining exercise interventions with other therapeutic strategies to optimize metabolic control in individuals with T2DM.

## Data Availability

All data produced in the present study are available upon reasonable request to the authors

## Acknowledgements

Special thank you to the research participants for their time and effort and to Simran Baid, Alexa Bernard, Mingming Cui, James Ha, Emma Hagen, Amanda Howitt, Sydney Hughes, Jessica Kosticak, Zhen Lin, Gabriela Parma, Meelap Patel, Celene Philip, Michelle Pieper, Ethan Pizano, Zach Powrozek, Julia Silverman, and Katelin Yaldo for expert technical assistance in exercise training and data management.

---

**Supplemental Table 1.** Primary and secondary antibodies.

| Antibody | Company | Catalog Number | Dilution |
| --- | --- | --- | --- |
| RAGE | Abcam | ab3611 | 1:1000 |
| ADAM10 | CST | 14194 | 1:1000 |
| TLR2 | CST | 12276 | 1:1000 |
| TLR4 | CST | 38519 | 1:1250 |
| MMP2 | CST | 40994 | 1:1000 |
| MMP9 | CST | 13667 | 1:1000 |
| TIMP 1 | CST | 8946 | 1:1000 |
| TIMP 3 | CST | 5673 | 1:500 |
| IRDye 800CW<br>Donkey anti-Mouse<br>IgG Secondary<br>Antibody | LI-COR | 926-32212 | 1:10000 |
| IRDye 800CW<br>Donkey anti-Rabbit<br>IgG Secondary<br>Antibody | LI-COR | 926-32213 | 1:10000 |

Receptor for advanced glycation end products, RAGE; a disintegrin and metalloproteinase 10, ADAM10; Toll-like receptor, TLR; matrix metalloproteinase, MMP; tissue inhibitors of metalloproteinases (TIMPs).

